# UK and other SARS-CoV-2-Covariants - Simulation Modeling 70% Increase

**DOI:** 10.1101/2021.02.05.21251230

**Authors:** Ernie Chang, Kenneth A. Moselle

## Abstract

CovidSIMVL, an agent-based contagion-based viral transmission simulation tool, was employed to simulate the effects of viral agents of differing levels of infectivity. The constructs “Velocity” and “Increase in Velocity” were operationalized in terms of rates of transmission events over successive iterations (generations) in a set of CovidSIMVL trials. Treating 40-70% increase in velocity as a target, based on reports in the literature for the UK variant (*VUI 202012/01)*, the series of trials reported in the paper demonstrate the calibration of CovidSIMVL parameters to produce increases in transmission rates of 40-70% above a baseline value. A series of follow-up studies is proposed to evaluate three different possible explanations for reported increases in SARS-Cov2-2 infections that are being attributed to spread of the UK and other variants: (a) simulations where the inherent characteristics of the virus (infectivity) are varied (genomic studies); (b) simulations where the behaviour of agents is varied (e.g., movement within and between spaces) while inherent characteristics of the virus are held constant (behavioural studies); and (c) simulations where both inherent properties of the virus and the behaviour of agents are varying to “tease out” the interaction between biologically-based contributions to increased case counts, and contextual/behavioural contributions (epigenetic studies).

## BACKGROUND

### New SARS-CoV-2 Variant Under Investigation/Variant of Concern

#### The World Health Organization stated on 21 December 2020

*On 14 December 2020, authorities of the United Kingdom of Great Britain and Northern Ireland reported to WHO that a new SARS-CoV-2 variant was identified through viral genomic sequencing. This variant is referred to as SARS-CoV-2 VUI 202012/01 (Variant Under Investigation, year 2020, month 12, variant 01)*.^1^

#### Citing Public Health England,^2^ the WHO reported on 31 December 2020

*Initial analysis indicates that the variant may spread more readily between people. Investigations are ongoing to determine if this variant is associated with any changes in the severity of symptoms, antibody response or vaccine efficacy.^3^* In this document WHO also identifies risk for reinfection as an area for investigation.

#### As per the *Economist*,^4^ 21 December 2020, and in keeping with various reports on increased rates of transmission, we abstract the following

1. This new variant is reported to be “40% - 70% faster”.
2. It is thought to be more transmissible.
3. The new variant’s proportion among total positive tests is increasing.

#### Note however from Volz (2020)^*5*^

*Estimating the epidemiological fitness of individual genetic variants during an emerging pandemic presents multiple challenges. The recent origin of SARS-CoV-2 combined with a relatively low rate of evolution means global viral genetic diversity is low, and many methods for identifying positive selection will have low sensitivity. Evidence for positive selection at spike position 614 and other sites has been suggested by statistical models based on the rate ratio of nonsynonymous to synonymous substitutions (Pond, 2020).^6^ However, the detection of positive selection by such methods does not necessarily imply the mutation enhances transmissibility, and effects of individual mutations on transmissibility will generally be low (Mac Lean et al*., *2020).^7^*

### Questions re: new variants

The following points would benefit from clarification.

#### Question #1 – re

“40 - 70% faster” – what is happening more rapidly, what is being measured?

a. New cases per day – is this referring to an acceleration in the rate of new cases per day, or perhaps referring *only* to such an acceleration?
b. Changes in rate ratios - is “faster” referring to an increase in proportion of new variant test positive tests compared to earlier variants [#3 above] over successive measurement points?
c. Is this increase in transmission fitness referring to an increase in the average number of infections associated with any given Index Case (reproduction rate or R_0_), such that population rates are increasing?
d. Is this a decrease in time to first detection in the lab compared to standard SARS-CoV-2 incubations?
e. Is this a decrease in time to first symptoms in test animals compared to standard?
f. Is this a decrease in time found through contact tracing between contact and first appearance of symptoms or first detection of virus?

#### Question #2 – re

“more transmissible” – does this relate to speed of transmission or spread? In particular:

a. Reproduction number-related – did contact tracing permit a time-based relationship between time of contact and first appearance of symptoms among contacts to show that the intervals were smaller, or that more infections per known contact were found?^8^
b. Were that laboratory studies with animals under controlled conditions of standard physical spacing and movement, where the introduction of an infection transmitter led to higher numbers being infected in shorter periods of time?
c. If “faster” is referring to changes in rate ratios (test positives for new variants vs earlier variants), have “founder effects”^9^ been evaluated as a potential cause for the increase, over and above any possibly inherent increase in infectivity associated with the new variant. Stated in slightly different terms: has it been shown that the new variants confer a statistically measured competitive advantage such that they are pushing out earlier less competitive phenotypes?
d. If an increase in the proportion of test positives for the new variant is correlated with an increase in the confirmed case rates, are there any biological features of the clinical phenotypes associated with the new variant that could be causal, e.g., measures of viral shedding.^10^

### Questions/issues addressed in this paper

These are all important questions, and they can be related back to one broad question concerned with causality, which can be broken down into three parts:

#### Question

if there are increases in confirmed cases that appear to be correlated with increases in the proportion of test positives for the new variant, what is(are) the causally operative factor(s):

a. **Factor 1** – **a new phenotypic variant (biologically based)** – re: measured increases in case rates that are correlated with increases in the proportionate rate of test positives for the new UK variant [or other variants]. Are these a product of increased transmissibility that is inherent in the biological properties of the new variant?
b. **Factor 2** – **behavioural/contextual** - if such a correlation exists, is it a product of behaviour, e.g., increased social contact and reduced social distancing over time, possibly combined with timing, e.g., increase in travel of social mixing over the end-of- December holiday season?
c. **Factor 1 x Factor 2 – ’epigenetic’ causes** -if the correlation exists, could it reflect a joint contribution of biologically-based increased infectivity ***and*** changes in behaviour?

The objective of this report is to set out a methodology that can be used to generate simulations that are configured to speak directly to each of the three causal explanations set out above. Because the intent is to evaluate the potential for a particular agent-based methodology, rather than evaluate all three causal options, the report is limited to a set of simulations that only addresses the first hypothesized causal explanation – biologically-based enhanced transmission “fitness”). As such, this report should be treated as preparatory clarification for a more complete body of work that would cover all three of the issues/questions identified above.

Specifically, this report takes on two key questions/issues:

1. How can CovidSIMVL (an agent-based simulation modeling tool) be configured to reflect an increase in infectivity associated with the virus?
2. What kind of summary measures of “X% faster” can be generated from the outputs produced by CovidSIMVL simulations.?

## METHODS

### CovidSIMVL –agent-based simulations of contagion-based transmission as a function of biological and/or behavioural factors

A large fraction of the epidemiological studies that characterize spread within populations and make predictions about the future employ compartmental equation-based models.^11^ These epidemiological models take sets of events as their inputs (e.g., daily new confirmed cases) and output sets of equations that generate curves that are best fits for the distribution of input values.

These models are suited to the task of predicting the future based on historical data, and within limits, they can enable inferences to be drawn about the impact that changes in behaviour or vaccinations or properties of a viral agent might have on future case rates. However, these classic epidemiological approaches are more limited when the intention is to determine from historical data the causal factors that govern dynamics that generate the events that these equation-based approaches model.^12^

Agent-based models of contagion-based spread do not start with events. They start with models of the dynamics that govern transmission and *produce* events – new infections – as outputs. By varying the conditions that govern the dynamics, it becomes possible to run virtual clinical trials or experiments. These agent-based approaches are inherently well suited to the task of evaluating questions related to causality – at least under those conditions where when ethical considerations, time or logistics preclude the possibility of running real clinical trials or completing bench science studies to evaluate causal hypotheses.

Because our intent is to provide a simulation engine that can be used to evaluate causal hypotheses relating to “faster spread”, we have employed CovidSIMVL.^13,14,15^. This tool (github.com/ecsendmail/MultiverseContagion), like other agent-based modeling tools, generates events by modeling the dynamics that determine whether/when those events occur within a context. Local contextual realism is injected into the simulations by configuring parameters to reflect those real-world factors (e.g., vaccine deployment; social distancing) that govern the emergence of events.

Note that unlike agent-based models that *assume* a value of R_0_ (reproduction number)^16^ and generate sets of events that are conditioned by that assumed value, CovidSIMVL treats R_0_ as an *emergent* characteristic of the properties of the virus and the properties of people in local contexts that are the real-world locus of any instance of transmission. It is aggregated across agents and computed at the end of a trial, rather than set at the beginning of a trial.

For details on how CovidSIMVL generates transmission events as outputs, based on values assigned to parameters that govern transmission, without assuming a reproduction number see ***Appendix I***.

### Parameters that can be adjusted in CovidSIMVL to reflect dynamics governing transmission

CovidSIMVL captures the dynamics governing viral transmission by setting parameters that fall within three domains:

1. <u>Viral-based temporal dynamics</u> -determinants of viral transmission that are keyed to viral load. These include incubation period, pre-symptomatic and symptomatic durations. Because viral transmission is dependent both on viral load, mechanism of transmission and proximity of agents, each agent is associated with what CovidSIMVL refers to as a “HazardRadius” or “HzR”. When the HzR of two agents overlaps, transmission can take place.
2. <u>Movement within local contexts</u> -movement of persons within a CovidSIMVL delimited space (a “universe”) determines whether/when the HzR of agents overlap. The movements of agents may be different across different modeled locations, such as in a pub, or a place of work such as a long-term care facility. As well, movement of different agents within a location may be different. These are captured by the CovidSIMVL “MingleFactor” or “mF” parameter. In any iteration (generation) within a given trial, mF varies stochastically from one generation to another according to a Pareto-like distribution of movements.
3. <u>Movement of agents from one location to another</u> (e.g., from home to a pub to home to a pace of work such as a long-term care facility). These moves are determined by a schedule that reflects typical movements of people in the real world over the course of time, e.g., a day and/or a week. The schedule may assign a different mF to different agents within and across universes, such that any given universe may include agents moving differently (e.g., patients *vs* staff in a long-term care facility).

These parameters collectively constitute an operational specification of the “dynamics” governing transmission within a given simulation trial. These are the causal factors.

### Setting HazardRadius Parameter in Simulations Concerned with Different Degrees of Viral Infectivity

Values for the parameters reflecting degree of viral infectivity and temporal dynamics are derived from the model of He and associates.^17^ This model (see ***Figure 1***) sets working reference ranges for viral loads and days after infection for incubation, pre-symptomatic transmission, first appearance of symptoms, and viral load tapering to below measurement thresholds in mild cases.

**Figure 1.**
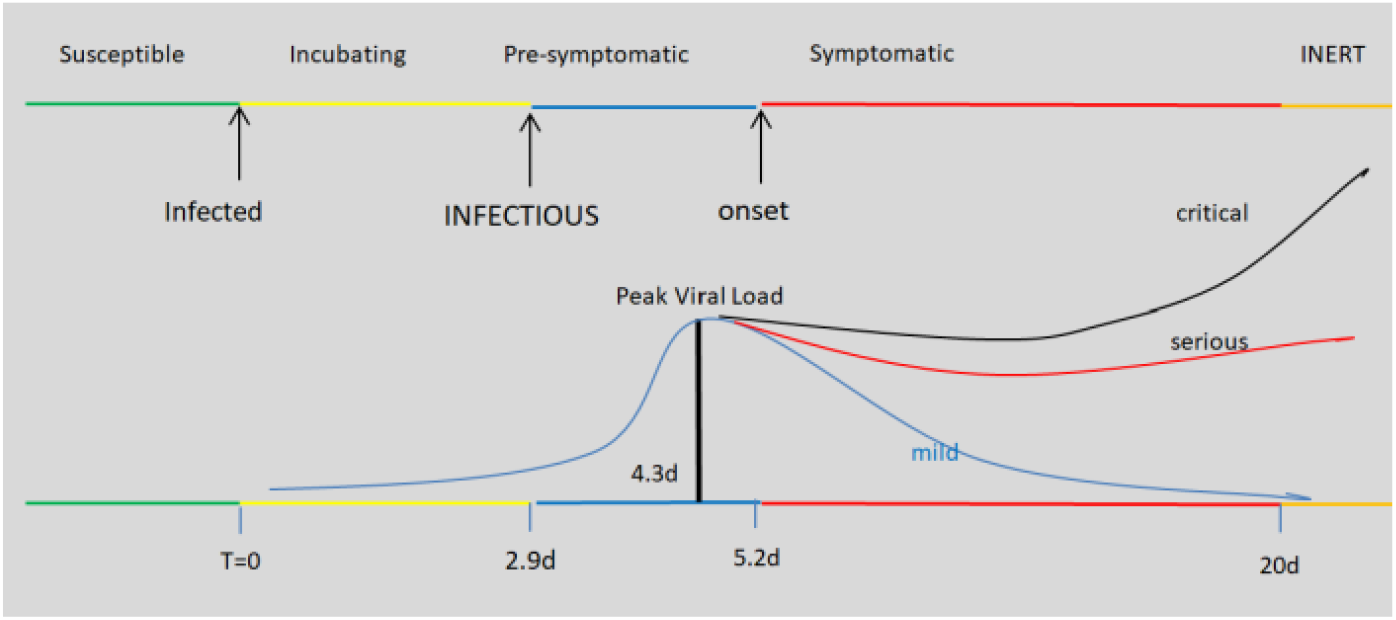
Configuring Primary Rules – Temporal Dynamics in Viral Shedding.

### Running CovidSIMVL trials to simulate different degrees of infectivity

A given CovidSIMVL trial is initiated by setting parameters, including HazardRadius (HzR) and MingleFactor (mF), both of which are given scalar values. *Changing values on HzR has the effect of changing the simulated degree of infectivity*.

The standard Out-of-Box settings for CovidSIMVL are for a population of 100 agents, with HzR of 5, in a fixed arena of 800×600, an mF of 1, and viral settings conforming to the Xi model as depicted in ***Figure 1***, above. This is treated as a control or a reference point, against which changes in the velocity of the infectious spread as a function of changes in HzR (see Metrics, below) can be evaluated.

For the results reported below, HzR was varied from a value of 5 to a value of 12. The other parameters were unchanged.

Trials are run until either one of two conditions are met:

1. There are no more Susceptibles, or no more Infective transmitters – the outbreak has self-extinguished. Number of generations required for this to occur is a marker of the “efficiency” or velocity of the epidemic spread, which is related to the infectivity of the virus and the behaviour of susceptible or infected agents within contexts. See Chang & Moselle for details.^18^
2. A specified portion of the original set of 100 susceptibles has become infected. Again, number of generations required to meet these benchmarks is a marker of the velocity of epidemic of spread.

For the results reported below, three benchmarks were used: 25% (i.e., 25 of 100 Susceptibles infected), 50% (50 of the Susceptibles infected) and 75% (75 of the Susceptibles).

### Metrics – Velocity; Increase in Velocity

#### Velocity

We measured the number of generations taken to reach criteria of 25 infections, 50 infections and 75 infections. We calculated the <u>Velocity</u> as follows:

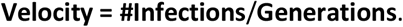

The **Generation** count is a measure of time. The **#Infections** is the number of Susceptibles (out of 100 at initiation of the trial) who have become infected when the criterion is reached.

The Velocity in the Out-of-Box configuration was calculated to serve as a control, i.e., as a denominator in measures of changes in velocity. Velocities were also calculated for different values of HzR.

#### Changes in Velocity

The Increase in Velocity is expressed as a percentage increase in Velocity relative to the Velocity for HzR= 5 in the Out-of-Box configuration. For each value of HzR, this change measure is computed at each of three points in the course of an outbreak – the points when 25%, 50% or 75% of the original Susceptibles have become infected..

#### Reproduction Number (R_0_)

R_0_’s were recorded in the CovidSIMVL console.log for each agent when the when a given trial terminated. For all of the trials with a HzR > 5, the trials terminated when all Susceptibles had become infected. R_0_ in ***Table 1*** represents the average number of transmissions by infected agents at the time of termination of the trial.

**Table 1.** Changes in Velocity of Spread as a Function of Changes in Infectivity (HazardRadius)

| HzR | Total Gen | R0 | Gens to Criterion |  |  | Velocity (#Infections/Gen)* |  |  | % Increase Velocity** |  |  |
| --- | --- | --- | --- | --- | --- | --- | --- | --- | --- | --- | --- |
|  |  |  | Gen to 25 | Gen to 50 | Gen to 75 | V.25 | V.50 | V.75 | 0.25% | 0.50% | 0.75% |
| 5 | 1190 | 2.02 | 306 | 442 | 538 | 0.082 | 0.113 | 0.139 |  |  |  |
| 6 | 1034 | 2.48 | 287 | 404 | 535 | 0.087 | 0.124 | 0.140 | 6.6% | 9.4% | 0.6% |
| 7 | 717 | 3.00 | 227 | 289 | 341 | 0.111 | 0.172 | 0.215 | 36.2% | 51.9% | 54.1% |
| 7.5 | 801 | 2.73 | 212 | 288 | 359 | 0.118 | 0.174 | 0.209 | 44.3% | 53.5% | 49.9% |
| 8 | 763 | 3.38 | 182 | 238 | 295 | 0.139 | 0.211 | 0.255 | 69.7% | 86.6% | 83.2% |
| 9 | 653 | 3.36 | 180 | 217 | 282 | 0.139 | 0.234 | 0.267 | 69.8% | 106.5% | 91.3% |
| 10 | 566 | 3.81 | 137 | 175 | 223 | 0.182 | 0.286 | 0.336 | 123.4% | 152.6% | 141.3% |
| 11 | 616 | 4.21 | 126 | 164 | 217 | 0.198 | 0.305 | 0.346 | 142.9% | 169.5% | 147.9% |
| 12 | 588 | 4.86 | 121 | 148 | 178 | 0.207 | 0.338 | 0.421 | 152.9% | 198.6% | 202.2% |

\*Generations to Criterion
\*\* Increase in Velocity relative to HzR = 5 Velocity as a reference standard.

## RESULTS

When HzR was increased from 5 to 12, the results in ***Table 1*** were obtained.

The top line represents a control condition based on Out-of-Box parameters. The number of iterations required to reach 25 or 50 or 75 infections are found under the “Gens to Criterion” headings. For a HzR set to 5, 1190 Generations were required for the trial to terminate. In this case when there were 2 remaining Susceptibles but no remaining Infectives.

R_0_ is counted and computed by tracking and recording all agents that have infected any Susceptible, and by counting the number of agents who become infected, so at the termination of a trial R_0_ for that synthetic epidemic is the total number of susceptibles infected/number of infecting agents.

R_0_ for the HzR = 5 trial is 2.02.

The velocities with which these criteria are attained are found by dividing number of Infected agents at the point that a given criterion is reached by the number of Generations required to reach that criterion. For example, V.25 is 25/306 = 0.08170 for HzR of 5, and V.50 is 50/442 = 0.113122.

The % Increase Velocity (“faster than”) is found from comparing the next trial(s) to the baseline HzR=5.

Thus, for HzR of 6, the velocity at V.25 is 25/287 = 0.08771, and calculating the gain from V.25#HzR5 is:

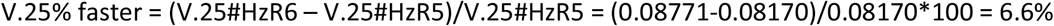

We can see that as the HzR increases, the Generation time decreases for increasing value of HzR, and that the Velocity gain increases correspondingly.

Regarding the key question: when is the new variant faster than the original by 40-70%? The simulation results from trial to trial vary slightly depending on when the measurements are taken (e.g., at 25% infected, 50% infected, 75% infected), because there are saturation effects toward the end of a trial, as Susceptibles diminish in supply. Overall, the fit for 40-70% *increase* is at HzR between 7.5 to 8, compared to the base line value of 5.

The following chart summarizes the above, visually.

**Figure 2.**
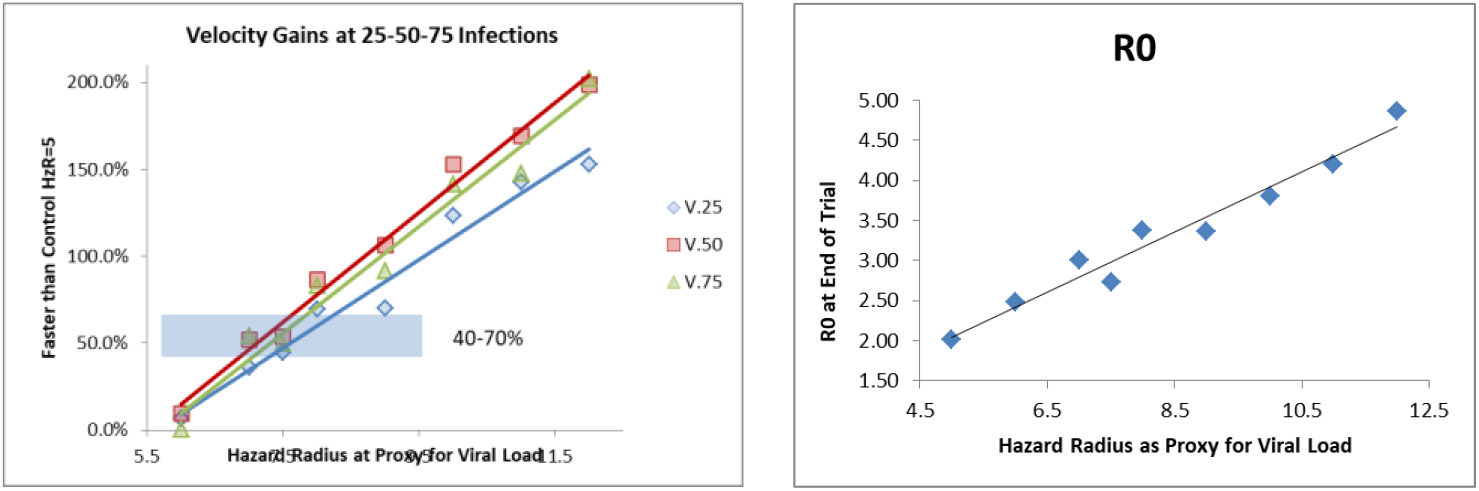
Velocity gains as a function of increase in HazardRadius (HzR).

## DISCUSSION

### 40% to 70% increase in Velocity

The simulated outbreaks reported in this paper show that the 70% faster can be achieved if the viral replication in-host (as captured by the HzR parameter) is sufficiently large, corresponding to HzR increase from 5 to 7.

As 5 is the radius, the volume increase is the cube of the radius given the volume of a sphere being 4/3 *∗ πr*^3^. Thus, for an increase in Velocity (as measured in this paper) reaching 70%, the viral load from the Xi model would have to increase proportionate to the volume increase from 5 to 7. Given that 5^3^=125 and 7^3^ is 346, this implies that the viral replication is roughly 2.5 times the currently prevalent strains of SARS-Cov-2.

We may consider a more purely spatial perspective, concerned more with the mechanics of transmission. In this case we look at the radius of a hazard “circle” rather than a hazard “sphere” that surrounds agents, e.g., assuming that transmission occurs roughly laterally *via* droplet transmission from one agent to another. These circles must overlap for viral transmission to occur. In this case, we would be looking at an increase from 5 to the second power to 7 to the second power, i.e., from 25 to 49, which is only a factor of close to 2.

We tried a number of other combinations of factors to bring about a 70% increase in the speed of infection, as we defined operationally above. For example, we decreased the incubation period of the Xi model (2.9 days) without significant effect, unless the viral replication was increased significantly, as in these reported trials. Results of those additional trials are available from the corresponding author.

### Reproduction numbers in CovidSIMVL, and relationship of findings to “real world” contexts

R_0_ for the HzR trial run with Out-of-Box parameter settings is 2.02. This does not necessarily mean that this trial represents the variant of SARS-CoV-2 that was spreading before the emergence of the UK or other new more infective variants. What it does mean is that the parameters for CovidSIMVL were set to capture dynamics (viral load, timings, agent movement) in such a way that changes in the number of infections over time produced a curve that reflected real world outbreak settings with a reproduction rate of approximately 2.00. This only means the simulation bears some dynamic relationship to real-world outbreaks in settings where the measured R_0_ is approximately 2.0. That could be settings where the original variant of SARS-CoV-2 was responsible for test positives. It could be a setting where the UK or South African or other variants predominate, but other controls (e.g., social distancing, contact tracing and isolation) were moderating the rate of spread.

The reproduction number, in and of itself, cannot say what is the biological phenotype responsible for the reproduction number unless there exists prior information that maps reproduction numbers onto distinct variants ***and*** it can be shown that other factors are not influencing the measured reproduction number.

### Next steps

CovidSIMVL is a simulation tool which supports situational analyses. It is not intended to be a predictive tool for IRL (“in real life”) epidemics. However, it is intended to reflect the dynamic interaction among factors that determine emergent characteristics of real-world processes, such as the emergence of contagious infections in local contexts and the spread of infection across functionally linked contexts *via* the interaction of biological properties of a viral agent, decisions and associated behavioural characteristics of persons (human agents), and their interaction in various real-world settings (e.g., school, home, work, recreation) within which their social and economic and personal lives unfold.^19^ CovidSIMVL supplies a sandbox in which experiments (including virtual clinical trials) can be undertaken that would be impossible to conduct IRL, given ethical or time constraints, and given the logistical challenges around mounting a real-world experiment in which several factors may be varying and need to be controlled *via* appropriate research designs.

In this paper, we report the results of one set of “experiments” in which we varied inherent infectivity of the virus. This employed one standard configuration for CovidSIMVL – high HzR, low mF. In a subsequent round of simulations we are evaluating the hypothesis that “faster spread” in ranges determined by new variants could be produced by behavioural change, holding degree of infectivity constant. To evaluate this hypothesized causal mechanism, we employ a second configuration paradigm in CovidSIMVL, in which HzR is set to a low value (and held constant) while mF is allowed to vary, until the percent increase in Velocity falls within a targeted range.

Building on previous work,^20^ we will also run a hybrid set of trials keyed to a hypothesized ’epigenetic’ set of causal mechanisms, creating a link between genotype and phenotype, where both Hzr and mF are modified, to determine different combinations of values on those parameters that could produce “faster spread”. The spatio-temporal framework within which transmission dynamics unfold in CovidSIMVL supplies a tailor-made ’epigenetic landscape’^21^ in which to observe these dynamics.

These additional trials are of more than academic interest. As Chang & Moselle^22^ show, these two different basic configurations – High HzR, Low mF, and Low HzR and High mF, produce quite different dynamics of spread over time, even if the endpoint (number of infected agents) is the same.

In particular, High HzR and Low mF produces the type of curve depicted in ***Figure 3***. This curve may be described as a wave that builds until it reaches a fixed amplitude, sweeping through a population of susceptibles. By sharp contrast, Low HzR and High mF produces the type of growth curve depicted in ***Figure 4***, which looks more like the usual exponential growth curves that describe random transmissions from a large spatially distributed array of infected persons.

**Figure 3.**
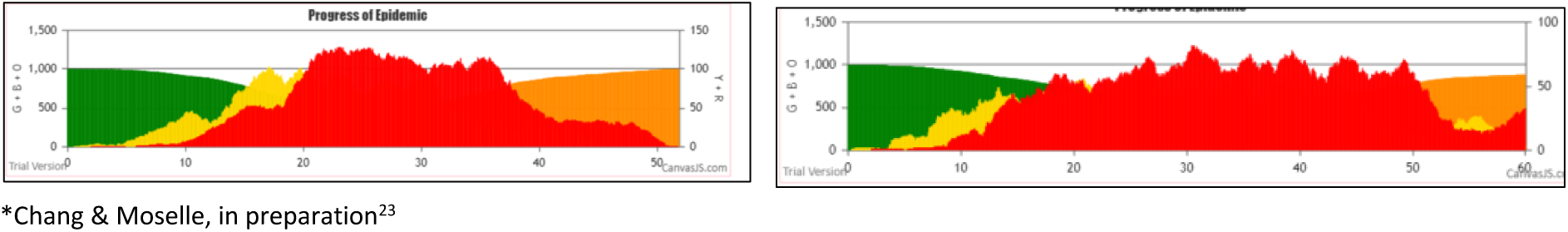
High HazardRadius (HzR), Low MingleFactor (mF)

**Figure 4.**
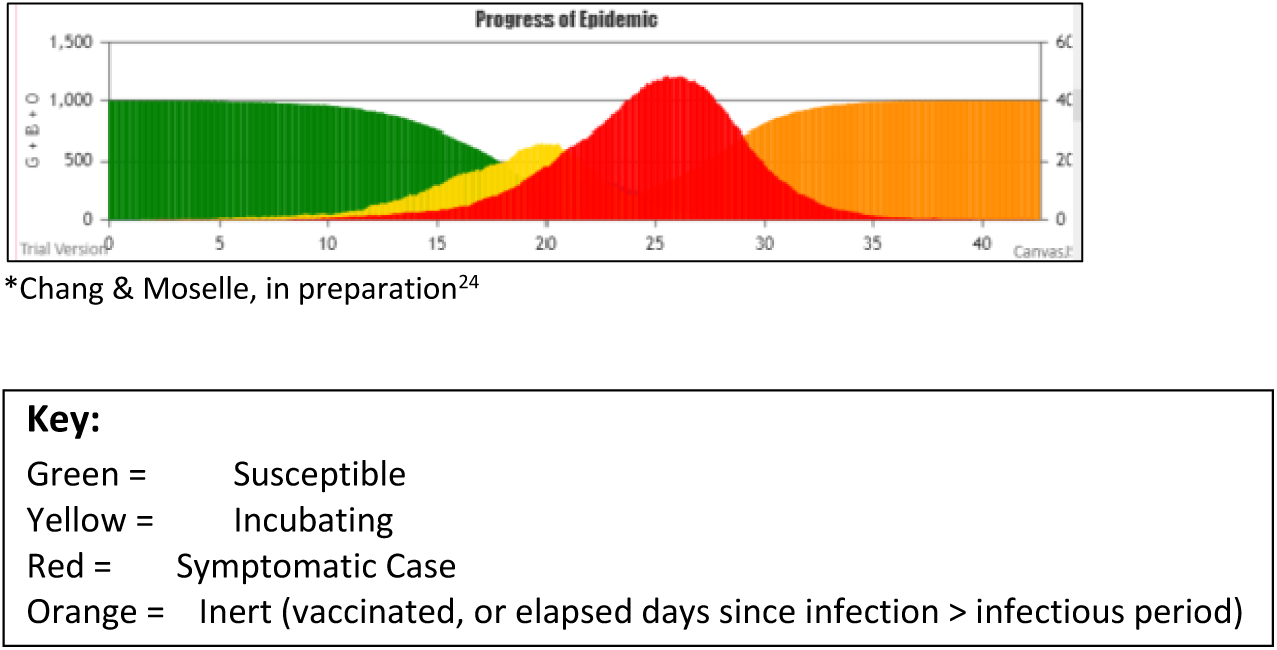
Low HazardRadius (HzR), High MingleFactor (mF)

The onset of rapidly increasing spread, and the rates at which the infection spreads in the middle and latter portions of the outbreak, are quite different in these two different simulation process models. This carries significant implications for expected impacts of different behavioural protections, depending on where and when they are put in place. It also carries strong implications for the manner in which vaccination strategies and schedules will confer protection. Different timing and levels of spread or protection can be expected when different vaccination policies are implemented in contexts where different causal mechanisms are determining the timing and the level of spread. Expected vaccination outcomes associated with the two different dynamics illustrated in ***Figures 3*** and ***4*** is the current focus of our work with CovidSIMVL.

## Data Availability

The work reported in the paper consists of simulated clinical trials, using CovidSIMVL - an agent-based modeling tool, available at github.com/ecsendmail/ MultiverseContagion.  The parameters used to generate the simulations are set out in the paper.

https://github.com/ecsendmail/MultiverseContagion

## Appendix

### How Does CovidSIMVL Work?

CovidSIMVL accomplishes the task of simulating viral transmission without assuming a reproduction number by employing a physical analogy of contagion among agents that move within one or more delimited spaces:

1. At any point in time an agent can be in one of four states: (a) Susceptible; (b) Incubating; (c) Symptomatic; or (d) Inert (no longer susceptible or infectious).
2. Transmission takes place when a person expresses a quantity of a virus sufficient to infect another agent. As such, typical changes in viral load over the course of an infection within a person determine whether transmission is possible or likely.
3. In CovidSIMVL, agents are located in a physical space and must be sufficiently close to one another that transmission can take place *via* mechanisms such as droplet transmission. In CovidSIMVL, all spaces (“universes”) have finite dimensions, so that the number of agents included in any given simulation will determine density of agents within the space.
4. Agents move within a space in characteristic ways, depending on their role or function/purpose. For example, staff in a long-term care facility move differently from patients.
5. The velocity at which people move within a space is characteristic of their role or function/purpose. Staff in long-term care typically move at a more rapid pace than patients.
6. People move across spaces in characteristic ways, depending on their role or function. For example, staff in a maximum 23 hour respite centre for homeless persons will work in that facility and also go home. Users of that facility will remain there for a maximum of 23 hours and then cross over into some street location, such as a soup kitchen.

Viral load has the effect of “wrapping” an envelope around a physical person or agent. The radius of this circle or agent is a reflection of two factors:

1. The mechanism of transmission (e.g. droplet transmission), and
2. The size of the circle. This is proportion to the viral load of a given agent at a particular point in the course of a simulation. In CovidSIMVL, we refer to this envelope as the “HazardRadius”) and it can be varied to reflect differences in infectivity of a virus. It also changes to reflect temporal dynamics of the virus, i.e., expected changes in viral load within a person over time.

In CovidSIMVL, the state of an agent changes from Susceptible to Incubating when the “wrapping” of a Susceptible agent overlaps with the “wrapping” of an infectious agent. When agents are moving over the course of multiple iterations (“generations”) in a simulation, viral transmission takes place when agents move into close proximity, where “closeness” is determine by the value of the HazardRadius.

CovidSIMVL keeps a record of the movement of agents when those movements give risk to transmission. In other words, CovidSIMVL models *dynamics* of viral transmission, in such a way that the reproduction number is a function of the temporal dynamics of the virus and the movement of agents in one or more spaces.

People move across spaces in characteristic ways, depending on their role or function. For example, staff working in a max 23 hour respite centre for homeless persons will work in that facility and also go home. Users of that facility will remain there for a maximum of 23 hours and then cross over into some street location, such as a soup kitchen.

### CovidSIMVL Trials

A trial consists of a series of iterations (Generations). With each iteration, agents are repositioned, and resulting proximity and simulated viral load determine whether transmission takes place. By varying some parameters (e.g., those that reflect inherent biological characteristics of the infective agent) while holding others constant, it comes possible to study the effect of changes to the different parameters.

The purpose of these experiments is not to predict a real-world state of affairs. The purpose is to generate “what-if” scenarios that reflect real-world contexts and dynamics, and evaluate the effects of changes to those dynamics via parameters. These changes fall within five broad classes:

1. Changes to the inherent properties of the virus, e.g., infectivity, and biologically determined temporal dynamics, expressed as changes in viral load within a given person over the course of the infectious illness.
2. Changes in the state of agents. For example, after a specified interval, e.g., 14 days, the state of vaccinated agent could change from Susceptible to Inert, as in not capable of being infected. Because interventions such as vaccinations do not confer protection on 100% of vaccinated parties, there are parameters within CovidSIMVL that can be adjusted to determine what proportion of a total simulation population are protected by a given vaccine at a given point in time.
3. Changes to the density of agents within a space.
4. Changes to the movement of agents within a space.
5. Changes in the movement of agents across spaces.

Collectively, parameters associated with these change can be used to simulate clinical trials or behavioural interventions.

